# Improved predictive diagnosis of diabetic macular edema based on hybrid models: an observational study

**DOI:** 10.1101/2023.04.05.23288182

**Authors:** JA Hughes-Cano, H Quiroz-Mercado, LF Hernández-Zimbrón, R García-Franco, JF Rubio Mijangos, E López-Star, M García-Roa, VC Lansingh, U Olivares-Pinto, SC Thébault

## Abstract

Diabetic Macular Edema (DME) is the most common sight-threatening complication of type 2 diabetes. Our goal was to develop an alternative method to optical coherence tomography (OCT) for DME diagnosis by introducing spectral information derived from spontaneous electroretinogram (ERG) signals as a single input or combined with eye fundus. To this end, an observational study was completed (n = 233 participants). Basal ERGs were used to generate scalograms and spectrograms via Wavelet and Fourier transforms, respectively. Using transfer learning, distinct Convolutional Neural Networks (CNN) were trained as classifiers for DME using OCT, scalogram, spectrogram, and fundus images. Input data were randomly split into training and test sets with a proportion of 80 % to 20 %, respectively. The top performers for each input type were selected, OpticNet-71 for OCT and DenseNet-201 for fundus and non-evoked ERG-derived scalograms, to generate a combined model by assigning different weights for each of the selected models. Model validation was performed using a dataset alien to the training phase of the models. None of the models powered by non-evoked ERG-derived input performed well. Metrics of the best hybrid models were all above 0.81 for fundus combined with non-evoked ERG-derived information; and above 0.85 for OCT combined with non-evoked ERG-derived scalogram images. These data show that the spontaneous ERG-based model improves all the performance metrics of the fundus and OCT-based models, with the exception of sensitivity for the OCT model, to predict DME. Combining non-evoked ERG with OCT represents an improvement to the existing OCT-based models, and combining non-evoked ERG with fundus is a reliable and economical alternative for the diagnosis of DME in underserved areas where OCT is unavailable.

**Author summary:** Providing an alternative diagnostic method to those that already exist for diabetic macular edema (DME) that is reliable and physically and economically accessible is needed in places where optical coherence tomography (OCT) is unavailable. In this work, we combined artificial intelligence (AI) classifying techniques with information from a newly introduced signal that can be captured in a non-invasive manner, the spontaneous oscillations of the electroretinogram (ERG). We found that if these signals alone are ineffective in diagnosing DME cases, they improve the performance of AI models based on either eye fundus or OCT in the prediction of DME. We therefore conclude that combining spontaneous ERG with fundus, which is a basic optometric test even in underserved areas, represents a reliable alternative to OCT for the diagnosis of DME. Also, combining OCT with spontaneous ERG signals will help ameliorate the diagnosis of DME.

## Introduction

Affecting 8.8 % of the world population and with an estimated number of cases reaching 783 million by 2045, type 2 diabetes is a modern pandemic (1,2). Diabetic macular edema (DME), characterized by the accumulation of exudative fluid in the macula, is the most common form of retinopathy that threatens vision in people with type 2 diabetes (3). DME is the leading cause of vision loss in diabetic individuals from 20 to 74 years-old (4). In Mexico, its prevalence has been estimated at 6.6 %, but most importantly, DME risk has been found to increase in the early stages of diabetes (5). As a major source of disability, it is, therefore, necessary to find alternative diagnostic methods to those already existing that are effective, accessible, and economical.

Artificial intelligence (AI) algorithms, particularly the start-of-the-art, continuously improving, machine learning techniques are generating enormous interest in diagnosing various diseases (6). In this context, AI algorithms known as convolutional neural networks (CNN) have been applied to the analysis of medical images, showing robust performance in the diagnosis and detection of conditions such as pulmonary tuberculosis from chest radiographs, malignant melanoma from skin photographs (6), and DME from optical coherence tomography (OCT) and fundus images (7). Even though biomicroscopy examination of the posterior pole of the eye remains the first step in DME diagnosis in many places, stereoscopic fundus photographs lack stereopsis, rendering the diagnosis unreliable (8). In contrast, OCT provides quantitative and qualitative biomarkers associated with visual and anatomical outcomes of DME (9), explaining why it largely supplanted fundus examination in DME diagnosis. However, OCT devices are the prerogative of rich countries.

Despite their good generalization features in various fields of medicine, CNNs come with a high computational cost and require large amounts of medical data for efficient model training (6,7). Transfer learning techniques address this issue by using pre-trained CNNs with millions of images of a different nature (e.g., ImageNet dataset (10)), transferring the weights obtained from this process, and training the network on a smaller dataset (6). Moreover, the development of hybrid models in AI (11), has enabled the use of different types of algorithms or different data sources with similar algorithms to generate more performant models (11,12).

In this study, we sought to improve the predictive power of fundus, as it is the most common DME test in the world. To this end, we took advantage of non-evoked or spontaneous electroretinogram (ERG) signals that have been recently shown to help predict risk factors of type 2 diabetes, including overweight, obesity, and metabolic syndrome (13). This is in view that ERG alterations are detected in patients with early DME (14,15), that portable devices can nowadays acquire ERG in a fully non-invasive way (16), and those 5-minute ERGs, in the absence of any light flash, that is in mock conditions, and under photopic conditions, which allows dispensing with mydriasis, is informative about early changes associated with diabetes (17).

We generated hybrid CNN algorithms powered by different types of data, including images of OCT, fundus, and non-evoked ERG spectrum obtained through either fast Fourier (spectrograms) or continuous wavelet (scalograms) transforms (13). Our data show that, although it is impossible to distinguish patients with or without DME using only non-evoked ERG spectral images, the performance of models using OCT and/or fundus images as input data can be improved when combined with spectral images of spontaneous ERG signals. Most notably, the hybrid model powered by fundus and mock ERG-derived wavelet scalogram images performs as well as the OCT one.

## Results

### Mock ERG-based models for the predictive diagnosis of DME

We first asked whether the mock ERG signals *per se* help predict DME cases. To this end, two CNNs, ResNet-50, and finely tuned DenseNet-201, powered by either the FFT or the wavelet-derived mock ERG power spectrum, were tested. **Figure 1A, B** shows the ROC curves and confusion matrices for CNNs based on FFT-derived information, while **Figure 1C, D** shows prediction results based on wavelet-derived information. The performance metrics of externally validated predictions are summarized in **Figure 1E**. For all models, the highest performance metric is specificity, and DenseNet-201 using scalograms is the most sensitive one (**Figure 1E**), but none of them showed performance metrics above 0.80.

**Figure 1.**
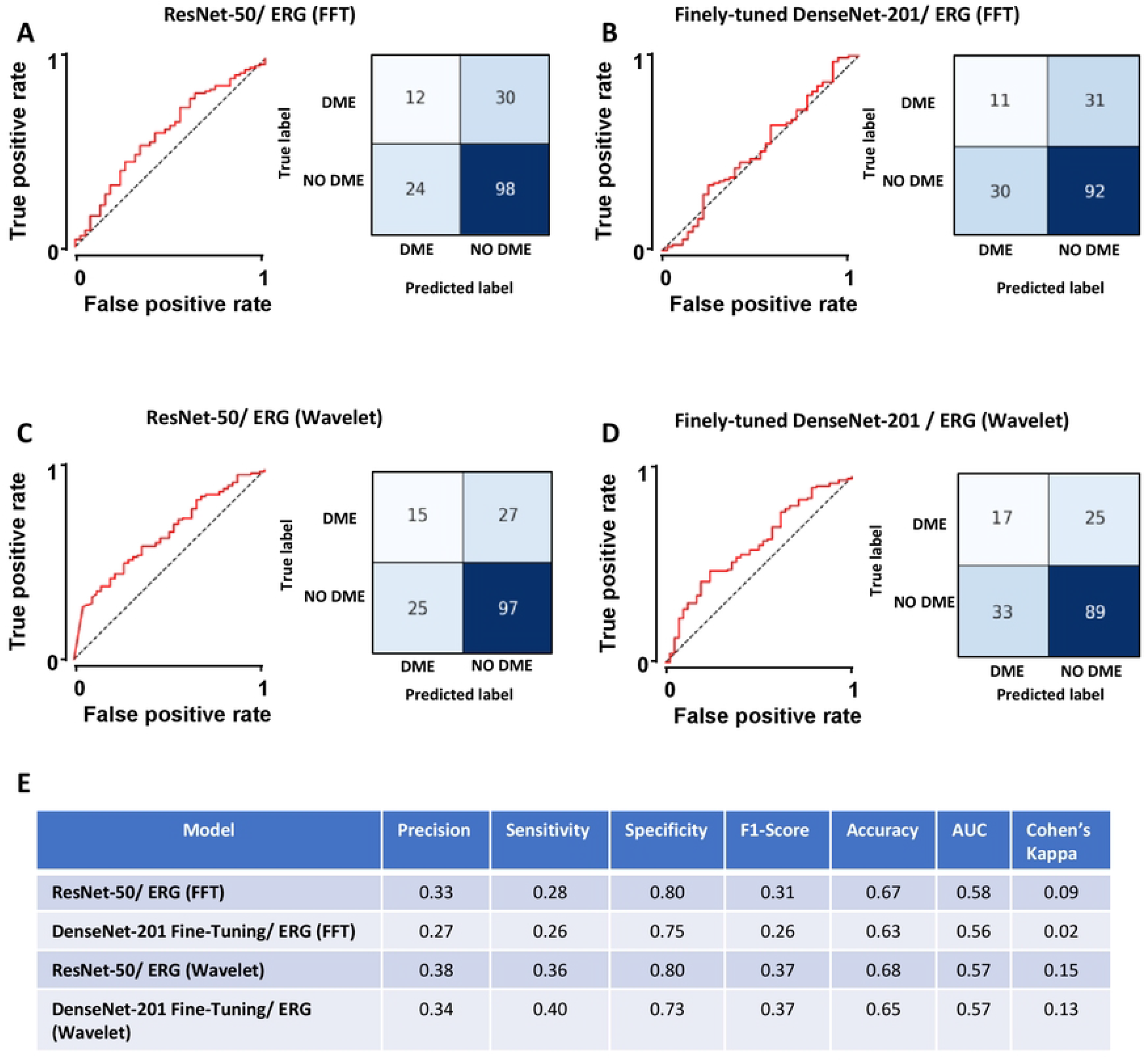
Diagram showing hybrid model operation. The best validated performing models were selected for each separate type of input (basal electroretinogram (ERG) scalogram, optical coherence tomography (OCT), and eye fundus images): OpticNet-71 for OCT and finely tuned DenseNet-201 for eye fundus and basal ERG scalograms. Predictions with the aforementioned models were done for each type of input (OCT, eye fundus, or scalogram images, n = 164 cases in total) and the result was multiplied by a scalar quantity *n*_*i*_that represents the adjustment percentage for each model. The three resulting values were then summed up to obtain a new prediction matrix, using the predictions made by each model. The maximum value (Max value) is obtained for each matrix row, the output being 1 with diabetic macular edema (DME) and 2 without DME.

In view of the poor performance of the models purely based on mock ERG-derived information to predict DME, we next analyzed whether the mock ERG transform could help improve the predictive performance of OCT and/or eye fundus images for DME.

### Hybrid models for the predictive diagnosis of DME

To implement hybrid models that combine mock ERG transforms, OCT, and eye fundus images, we first determined which models based on each of these entries independently were best, and then used them as inputs for the hybrid models.

As previously shown, finely-tuned DenseNet-201 trained with mock ERG-derived scalograms was the least bad in predicting DME (**Figure 1E**).

As for OCT-based models, external validation using 1,229 images from our database (**Table 1**) showed better results for OpticNet-71. Compared to DenseNet-201, its precision (0.93 vs. 0.52), sensitivity (0.93 vs. 0.91), specificity (0.99 vs. 0.83), F1-score (0.93 vs. 0.72), accuracy (0.98 vs. 0.84), ROC AUC (0.96 vs. 0.87), and Cohen’s Kappa (0.91 vs. 0.57) were largely higher (**S1 figure**, panels A, B, and E). Using the 164-image set (**Table 1**) of Srinivasan *et al*. (23), we further confirmed that the best overall performance for the predictive diagnosis of DME is for OpticNet-71, with metrics above 0.84 (**S1 figure**, panels C, D, and E). The sensitivity for DenseNet-201 was greater than the one of OpticNet-71 (0.95 vs. 0.93); nonetheless, the rest of the metrics were not (Precision: 0.85 vs. 0.63, specificity: 0.94 vs. 0.8, F1-score: 0.89 vs. 0.75, accuracy: 0.94 vs. 0.84, ROC AUC: 0.94 vs. 0.88, and Cohen’s Kappa: 0.84 vs. 0.64, respectively) (**S1 figure**, panels C, D, and E).

**Table 1.** Distribution and composition of the databases used. It is important to highlight that the images used for model training and testing were not used for validating the model performance. TO this end, independent images were used. Furthermore, the only datasets partially generated with data augmentation were those containing spectrograms and scalograms, but only for the DME group.

|  | EYE FUNDUS |  |  | OCT |  |  | SCALOGRAMS AND SPECTROGRAMS |  |  |
| --- | --- | --- | --- | --- | --- | --- | --- | --- | --- |
|  | Dataset | DME | No DME | Dataset | DME | No DME | Dataset | DME | No DME |
| **Training and Test** | Present study | 1,011 | 373 | Kermany's | 11,598 | 26,565 | Present study | 141 | 1,048 |
|  | Messidor | 0 | 151 | - | - | - | Present study with data augmentation | 846 | 1,048 |
|  | Pachade's | 0 | 180 | - | - | - | - | - | - |
|  | Giancardo's | 0 | 115 | - | - | - | - | - | - |
| **Validation** | Present study | 42 | 122 | Srinivasan's | 36 | 53 | Present study | 42 | 122 |
|  | - | - | - | Present study | 176 | 964 | - | - | - |

Three different models were tested for eye fundus image-based DME predictive diagnosis and externally validated with n = 164 (DME: 42, No DME: 122, **Table 1**). **S2 figure** (panel A) shows the ROC curve, confusion matrix, and performance metrics for the Resnet-50 model. At the same time, similar data are reported in **S2 figure** (panels B and C) for the finely tuned MobileNet-V2 and DenseNet-201 models, respectively. If ResNet-50 had a high sensitivity (0.86) and MobileNet-V2 a very high specificity (0.98), the DenseNet-201 model obtained overall more robust performance metrics, especially for F1-Score and Cohen’s Kappa that evaluate the performance of models trained with unbalanced classes (**S2 figure**, panel C).

Next, based on the demonstration that OpcticNet-71 trained with OCT images, DenseNet-201 trained with eye fundus images, and DenseNet-201 trained with scalograms were the best individual models, we created hybrid models based on the linear combination of each individual model prediction matrices (**Figure 2**). All possible combinations of weight factor values were systematically analyzed, considering incremental steps of 1 %. Heat maps for performance metrics of all hybrid models are shown in **Figure 3**. For the hybrid model that combined both OCT and eye fundus images, the highest metrics (all above > 0.87) were obtained when the weighting factor n1 was 0.60 and the n2 of 0.40 (**Figure 3A**), meaning that the contribution of OCT images was of 60 %, while the one of fundus images was of 40 %. In these conditions, precision reached 0.90, sensitivity 0.90, specificity 0.97, F1-score 0.90, accuracy 0.95, ROC AUC 0.94, and Cohen’s Kappa 0.87.

**Figure 2.**
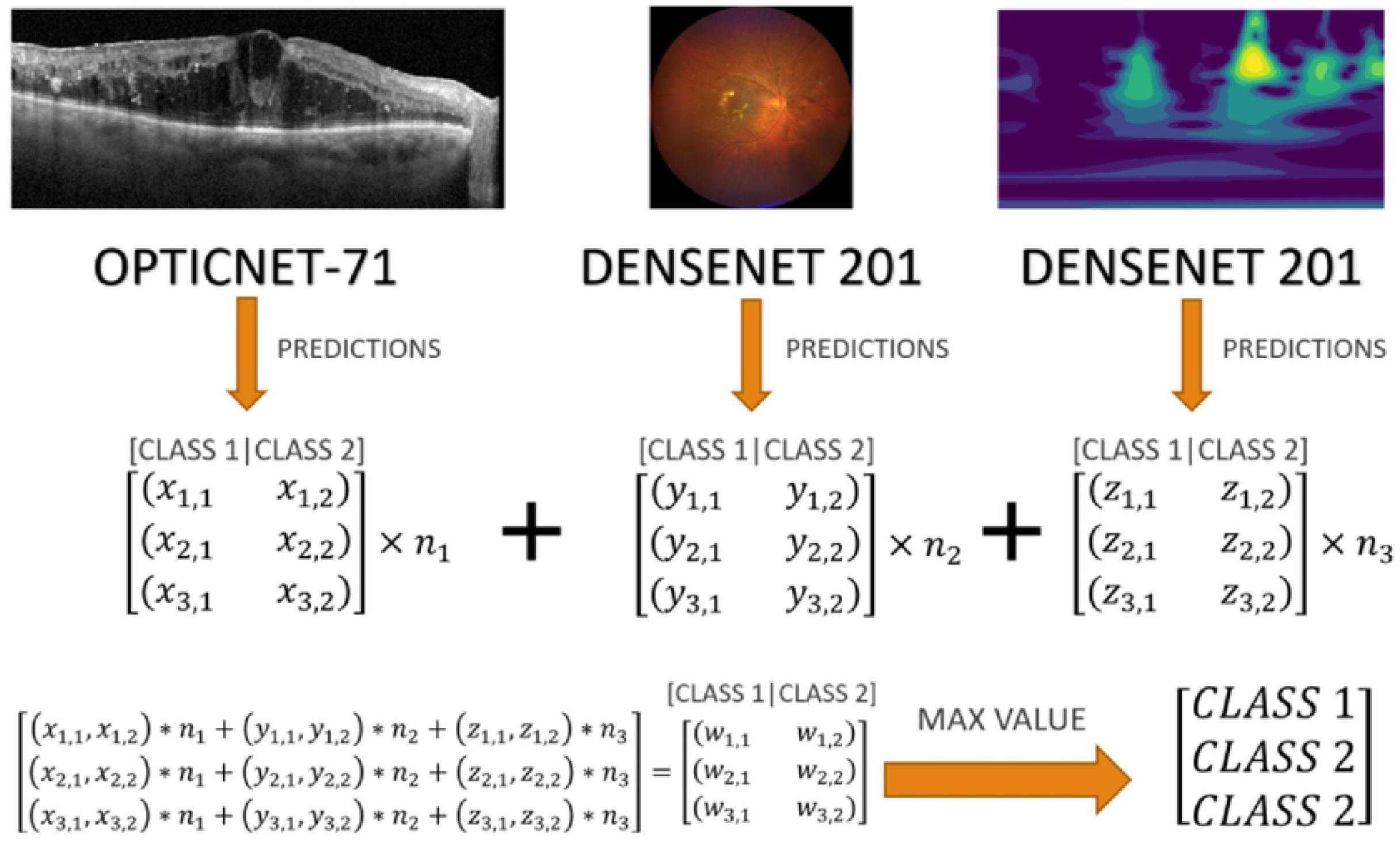
Performance of the spontaneous ERG-based models to predict DME. Receiver-Operating Characteristic (ROC) curves and confusion matrixes corresponding to the **(A)** ResNet-50 model fed with fast Fourier Transform (FFT)-derived spectrograms, **(B)** finely-tuned DenseNet-201 model fed with FFT-derived spectrograms, **(C)** ResNet-50 model fed with Wavelet Transform-derived scalograms, and **(D)** finely-tuned DenseNet-201 fed with Wavelet Transform-derived scalograms of spontaneous ERGs. The validation dataset included n = 42 cases with DME and n = 122 cases without DME. **(E)** Summary of all above-models’ performance metrics.

**Figure 3.**
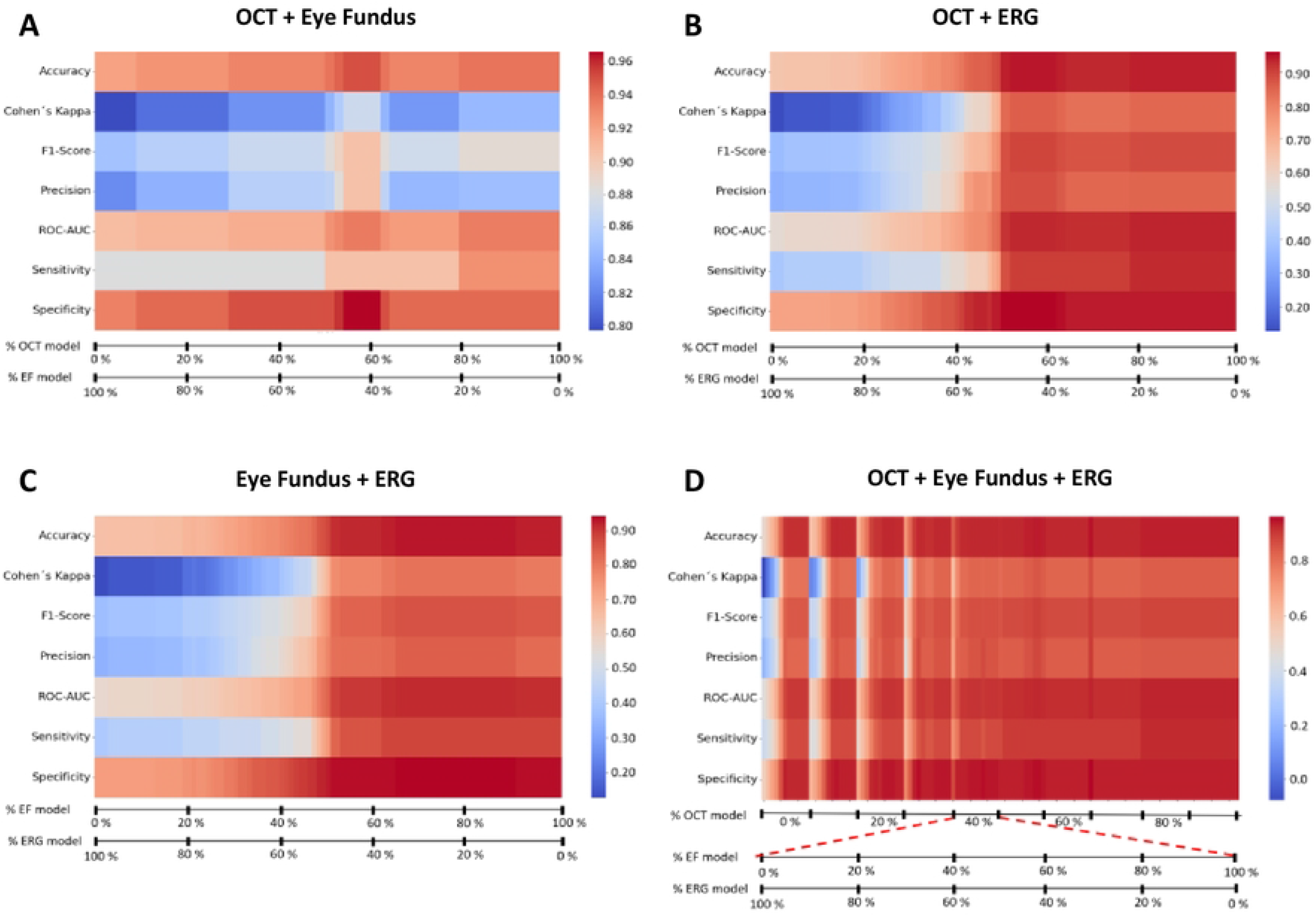
Optimization of hybrid models. Heatmaps showing the performance metrics of double and triple hybrid models, according to the proportion assigned to each used models, i.e. **(A)** OCT and eye fundus images, **(B)** OCT and basal ERG-derived scalogram images, **(C)** eye fundus and ERG-derived images, and **(D)** all three different types of data, as indicated by the percentages in the X-axis. All combinations summed up to 100 %. For the triple hybrid model (D), the remanent percentage of each 10 %-subdivision of the OCT-axis (e.g. 60 % in the case of 40 % assigned to OCT) is divided between eye fundus and ERG-derived images, as indicated in the two lower X-axes. Optimizers looked for the optimal *n*_1_,*n*_2_,*n*_3_values, as introduced in **Figure 2**.

For the hybrid model that combined both OCT and mock ERG-derived scalogram images, the model is optimal when the n value for OCT is 0.60 and for mock ERG 0.40 (**Figure 3B**), since that, in these conditions, precision reached 0.88, sensitivity 0.90, specificity 0.96, F1-score 0.89, accuracy 0.95, ROC AUC 0.93, and Cohen’s Kappa: 0.85. Most notably, these data show that including mock ERG-derived information improved the specificity, precision, Cohen’s Kappa, and accuracy of the OCT-based model (for reference, see **S1 figure**, panel A).

For the hybrid model that combined eye fundus and mock ERG-derived scalogram images, optimization occurred in a wide range of combinations from 50 % eye fundus and above and 50 % ERG and below, respectively (**Figure 3C**). For example, when 70 % of the input comes from the eye fundus-based prediction and 30 % from the mock ERG-based prediction, precision reached 0.84, sensitivity 0.88, specificity 0.94, F1-score 0.86, accuracy 0.93, ROC AUC 0.91, and Cohen’s Kappa: 0.81 (**Figure 3C**). All above 0.8 performance metrics of the DenseNet-201 model trained with eye fundus images (**S2 figure**, panel C) increased when it is combined with the mock ERG model in the 70-30 % proportions, except for sensitivity. We also created triple hybrid models that combined mock ERG, fundus, and OCT image inputs, and observed that several combinations led to optimal performance metrics (**Figure 3D**). The most notable optimization happened when n1 = 0.60, n2 = 0.40, and n3 = 0, corresponding to the double hybrid model that combined OCT and mock ERG-derived scalogram images (**Figure 3D**). Nonetheless, there are other optimized combinations, like the n1 = 0.60, n2 = 0.28, and n3 = 0.12, for OCT, eye fundus, and mock ERG-derived scalogram images, respectively, that reached a precision of 0.88, sensitivity of 0.90, specificity of 0.96, F1-score of 0.89, accuracy of 0.95, ROC AUC of 0.93, and Cohen’s Kappa of 0.85.

A single heat-map included all hybrid models’ performances (**Figure 4**).

**Figure 4.**
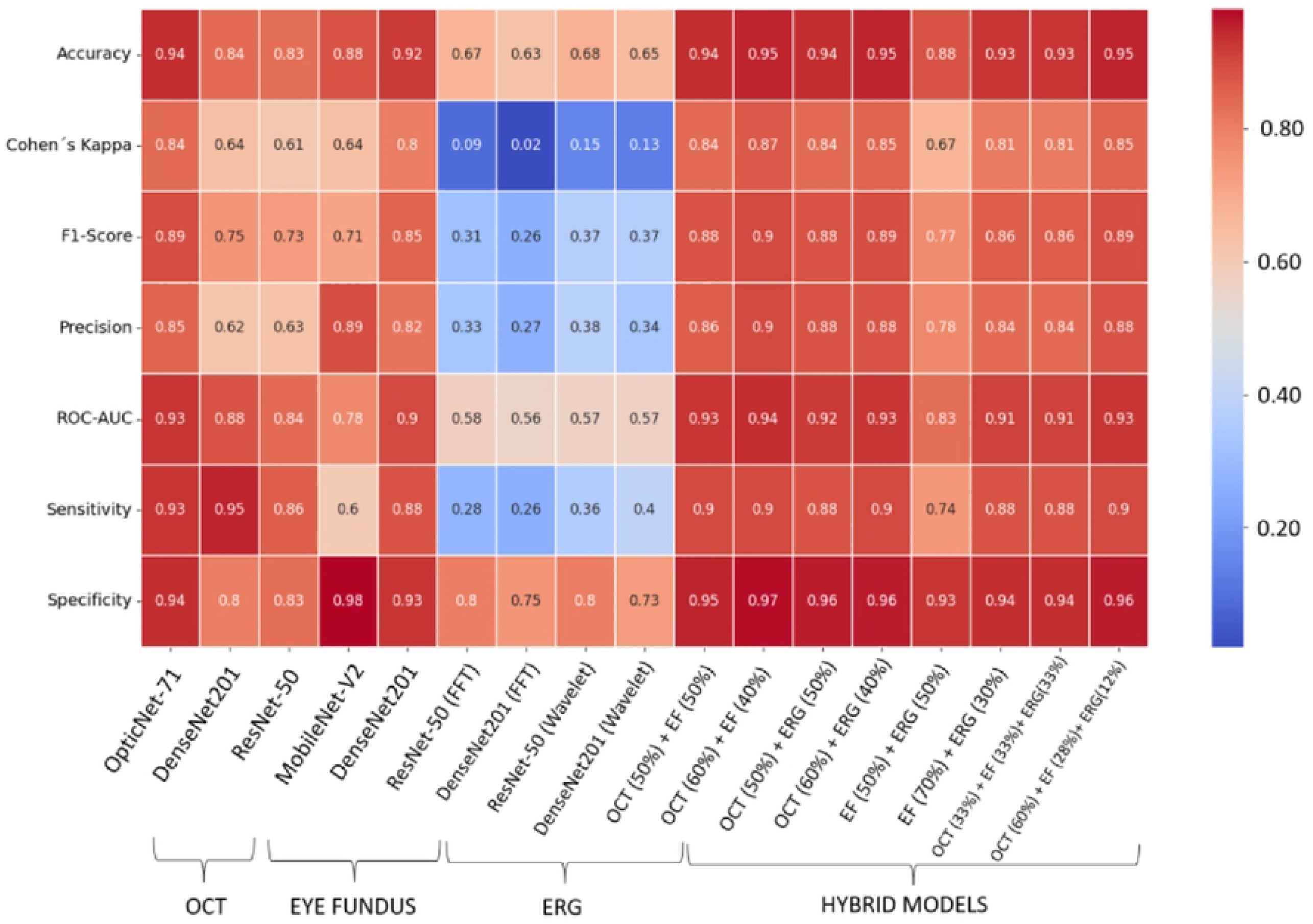
DME-predicting models’ comparison. Heatmap showing the performance metrics obtained for each tested models, including the ones based on one type of input (OCT, eye fundus or ERG-derived images), as well as double and triple hybrid models. External validation dataset: n = 42 for DME and n = 122 without DME.

## Discussion

DME continues to be the leading cause of preventable blindness in the working-age population in the world (8). Therefore, early detection programs are extremely important in treating this DR complication. Introducing new imaging modalities and technological advances has facilitated both early detection and follow-up of patients with DME, particularly OCT angiography and AI. However, OCT exams are not accessible to everyone in developing countries, due to their high cost and lack of equipment. In this context, we showed that a CNN model powered by images of changes in the frequency spectrum of basal ERG signals could not be used to diagnose DME, but that the performance of OCT and fundus image-based classifiers in predicting DME can be improved by combining them with the mock ERG transform cues.

The poor performance of our model based on mock-ERG signals alone can be explained by the state of the disease, the nature of the data, and the characteristics of the used models. At continuation, we will discuss each one of these issues.

DME is a complication of DR, it, therefore, develops in association with different degrees of DR, ranging from moderate non-proliferative to advanced proliferative stages (29). Furthermore, DR, classically defined as microangiopathy affecting the retinal vessels (30), is known to affect retinal neurons (31). Neuronal apoptosis begins with retinal ganglion cells but can also affect other retinal nerve cells, such as bipolar cells, amacrine cells, and photoreceptors (31). This neuronal damage can be evidenced by latency delays in multifocal ERG (32). Our training, test, and validation data contain ERG records from DME or non-DME patients diagnosed with DR to some degree. Therefore, spectral images from mock ERGs likely show similar changes in both DME and non-DME groups, confusing the models. We confirmed this scenario, since the performance metrics of the models trained to distinguish DR and no DR classes exceeded 0.60, unlike the values obtained for the validation of the DME class prediction, indicating that the concomitant presence of DR with DME takes part into the poor performance of our models.

Even though the models trained for DR classification performed better than those for DME, they are not robust enough to make a classification of sufficient quality for clinical use, which suggests that other variables, like the nature of the data and the models themselves, are influencing the classification.

If, to the best of our knowledge, there is no record of the relevance of basal ERG transform images for human disease prediction, previous works have reported the use of electrophysiological signals, such as EEG, transformed into spectral images to train CNNs to recognize original data from synthetic ones (33,34). As for this CNN model that performs relatively poorly (metrics of ∼0.6 for EEG-derived scalograms) (34), the time-frequency representations of our data may not express the necessary or sufficient information (67). Our sampling frequency is 2 kHz, but some data may be lost when converting the original signal to a spectral image. Spectrograms with too low spatial resolution have been shown to impair predictive model performance (35). Additionally, our dataset consisting of just over 1,000 images is very small compared to the 14 million image database usually used for CNN training (34). Data augmentation helped us partly circumvent this limitation, but expanding our dataset is necessary for the near future. Moreover, we dealt with the problem of missing data by using the transfer learning technique (10). However, this may have initialized the weights for detecting object characteristics that are not present in the spectrograms as proposed by Ruffini *et al*. (36), which may further explain why our methodology was inefficient in generating the DME diagnosis through images of mock ERG transforms. In addition, given the effectiveness of time series from spontaneous ERG oscillations in predicting risk factors for type 2 diabetes (17), the predictive value of time series data rather than images from mock ERG transforms in classifying DME and non-DME cases remains to be studied.

Our data showed that, when combined with the best mock-ERG model, the performance of the best OCT model to predict DME improved in terms of specificity, precision, Cohen’s Kappa, and accuracy. However, its sensitivity has decreased. Similar observations were found with the best fundus model, being more performant when combined with the best mock-ERG model, except for its sensitivity that remained unchanged.

It is recognized that mixed methods of prediction, which use multiple learning algorithms, improve the performance of predictions obtained by individual learning processes (37). As observed for analyzing sentiment in the text of low-resource languages (12) and for detecting esophageal cancer (38) by combining CNNs and support vector machine, our hybrid models provided better results than individual models to predict DME cases. It has been empirically verified that this improved performance relates to the combination of distinct model characteristics and the variability in data of different origins (24). We further believe that this improvement is partly due to the complementation of structural information with functional data (26). Structural images, like OCT and fundus, are most commonly used for retinopathy detection, including DME (39). These images are very useful when it comes to localize a lesion (40). However, their very nature gives them the intrinsic spatial resolution limit and the caregiver’s ability to interpret the image. In addition, they offer a static view of the retina. In contrast, ERG provides information on retinal activity, at a global or specific level (certain layers of the retina or cell type), depending on the stimulation protocol used. ERG can inform about subtle changes long before any structural alterations can be detected using images. Nevertheless, its application which consists of exposing the subject to a series of specific light flash protocols can be time-consuming, often requires dark adaptation and sometimes mydriasis, and is not recommended for DME detection (41), even though patients with DME showed alterations in some ERG parameters (42). In this context, the predictive power of 5-minute photopic ERGs in the absence of any light flash demonstrated by our data is a huge improvement. More particularly, scalograms obtained from the mock-ERG transforms can be considered functional images of the retina. Having both structural and functional information brings a more comprehensive view of the retinal tissue to the model, thereby improving its capacity to predict DME.

Another issue concerns the performance metrics of the models during training and testing. Accuracy and loss metrics were quite good for both the OCT and fundus models, but not for mock ERG-derived scalogram models. The large number of OCT images used for training largely explains the training performance of the OCT models (27,28). At the same time, the good quality and the transfer learning (43– 45) surely benefited the eye fundus models. In contrast, mock ERG-derived scalogram models were subjected to overfitting since the accuracy of the testing exceeded that of the training (46). The small amount of data available for training (141 spectral images) is likely responsible.

As previously mentioned, an interesting result is that, although the hybrid models generally performed better, the single models that use OCT images as input were the most sensitive. Thanks to the vast OCT image database (>30,000 images), the training was done, using 11,598 OCT images, thereby allowing the model to extract information from a larger number of diseased subjects, facilitating the recognition of true positives (28). In contrast, only 1,011 fundus images and 846 spectral images of mock-ERG after data augmentation were available for these respective model training, likely accounting for the higher sensitivity of the OCT models. We believe that when combining the models’ probability matrices, the mock-ERG and fundus models’ low sensitivities tended to decrease the OCT models’ high sensitivities. An alternate explanation is that unlike fundus images, which only show the inner face of the posterior part of the eyeball, allowing the observation of certain structures, such as the macula or the optic disc, and provides with some qualitative information, OCT images provide quantitative information, like the retinal layer thickness, as well as the presence or the absence of subretinal or intraretinal fluid (47). This translates into a greater amount of information about the retina, increasing the sensitivity of the prediction.

In our view, one of the most interesting results of this work is that the performance metrics of the combined fundus plus mock ERG-derived scalogram model in the 70%:30% proportion are comparable to that of the best OCT model. Both present Cohen’s Kappa greater than 0.8 and F1-score greater than 0.85, which, as previously mentioned, are the best metrics for evaluating models with unbalanced classes (28). That the sensitivity of the OCT model is superior to the one of the hybrid model (0.93 vs. 0.88) has been discussed above. These results become particularly interesting knowing that the cost of an OCT scanner varies widely from US$35,000 to $100,000 (48). In comparison, the cost of a non-mydriatic camera range from US$10,000 to $20,000 (82) and portatile electroretinographs can be as cheap as approximately US$4,800 (49). On the eve of being certified, we also know about other ERG prototypes that could even be cheaper. The purchase of the most expensive non-mydriatic camera in conjunction with the electroretinography gives an approximate cost of US$25,000, which remains cheaper than purchasing the most economical OCT device. In general terms, performing a fundus study plus ERG is cheaper than the OCT study. Thanks to our model, this information is now helpful for DME predictive diagnosis. This is relevant because eye care is inaccessible to many people (lack of specialists, insufficient infrastructure, and transportation to clinics) (50). Our work contributes to the view that AI-based diagnostic methods can solve the problem of the lack of specialists, particularly in the eye care area (50). Furthermore, our model deals with already trained models, minimizing the computational cost. It is interesting to note that our model can even be used on mobile devices (51), further reducing the use of resources.

In conclusion, using of the fundus-mock ERG hybrid model is viable and relevant for diagnosing of DME in current medical practice.

## Materials and Methods

### Ethics

The ethics committee for human participants of the Mexican Institute of Ophthalmology (IMO), the National Committee of Ethics (reference: CONBIOÉTICA-09-CEI-006-20170306), and the Research Committee at the Asociación Para Evitar la Ceguera (APEC, 17 CI 09 003 142) approved this study. Written informed consent was provided by all subjects. All procedures were done according to the principles of the Helsinki Declaration.

### Human data

Since we are introducing a completely new parameter, namely unevoked electroretinogram (ERG) signals, for the predictive diagnosis of diabetic macular edema (DME), our study serves as a pilot survey of the population and therefore we cannot determine the sample size (18). A total of 321 adult subjects aged between 30 and 80 years (mean: 48.13 ± 0.71 years, 165 females) with or without diabetes, were enrolled between February 26, 2015 and December 2019 and from September 2021 and December 12, 2023 in the IMO of Querétaro (mean age: 50.81 ± 1.57 years, 54 females) and between August 10, 2021 and March 20, 2022 in the Asociación Para Evitar la Ceguera (APEC) in Mexico City (mean age: 45.77 ± 1.20 years, 119 females). 233 (age mean: 44.31 ± 0.72 years, 118 females) completed all tests required for the current study.

Subjects underwent an anamnesis and an initial optometric examination to ensure that they were eligible to participate. The exclusion criteria were ages outside 30 to 80 range, lens opacity, myopia greater than 6 diopters, glaucoma or other concomitant ophthalmologic disorders, ocular anomalies (e.g., surgery, trauma), recent use of laser or anti-angiogenic intravitreal administration, and cornea problems that disable ERG recordings. All tests were performed as described in (13). Patient diagnosis for DME, diabetic retinopathy (DR), or other eye issues was established by experts at IMO (M.G.R., R.G.F., E.L.S.), APEC (H.Q. and L.F.H.Z.).

### Electroretinogram (ERG) signals

Non-evoked ERGs were registered using customized protocols with either RETIMAX (CSO), Moonpack (Metrovision), or RETeval (LKC Technologies) electroretinographs. Under light conditions (∼400 lux), the contour of the eye and the forehead of the subject were cleaned before placing both recording and reference electrodes. ERGs consisted of 5-minute recordings in the absence of any light flash under photopic conditions (400 lux). Recording conditions included a band-pass filter of 0.3 Hz to 1 kHz and an acquisition frequency of 2 kHz. Subsequently, raw data were digitally filtered between 0.3 and 40 Hz, as previously described (17). ERGs were then divided into one-minute segments to maximize the number of samples. From the ERGs of 233 patients (151 from APEC and 82 from IMO), 1,353 one-minute ERG fragments were obtained, 183 from patients with DME and 1,170 from patients without DME.

As part of this nascent research, we tested both Fourier and wavelet transforms. The Fourier transform captures frequencies that persist over an entire signal, which may not serve for signals with short intervals of characteristic oscillations (19). The wavelet transform is a good alternative because it decomposes a function into a set of wavelets (20). Spectrograms were obtained from each one-minute ERG fragment using Fast Fourier Transform (FFT) with the “Multitaper Spectrogram Code” tool (21). Similarly, scalograms were generated using the Scipy 1.7.1 & MatplotLib 3.5.1 libraries for Python 3.8.8.

### Image datasets

We used OCT images from three public datasets. Kermany’s dataset contains 11,598 images with DME and 26,565 without DME (22); Srivasan’s is formed by macula-centered OCT images of DME (n = 36) and non-DME (n = 53) patients (23), and our dataset (https://github.com/Traslational-Visual-Health-Laboratory) contains 1,140 OCT images: 176 images with DME and 964 without DME.

We used eye fundus images from four open sources, including the MESSIDOR (24), Pachade’s (25), Giancardo’s (26), and our own (https://github.com/Traslational-Visual-Health-Laboratory) datasets. Fundus images with no pathology were pooled (n = 151 from MESSIDOR, n = 180 from Pachade’s, n = 115 from Giancarlo’s, and n = 495 from ours), as well as fundus images with DME (n = 1,053, only from our dataset).

From the 1,353 non-evoked ERG-derived spectrogram and scalogram images, 164 were set apart for model validation: 42 with DME and 122 without DME. The remaining 141 ERG images with DME were subjected to data augmentation by modifying horizontal rotation, horizontal displacement, contrast, brightness variation, and adding salt and pepper noise, as previously described (27). After data augmentation, the non-evoked ERG-derived spectrogram and scalogram images dataset included 846 images with DME and 1,048 without DME.

Image dataset information is summarized in **Table 1**.

### Predictive models

Convolutional Neural Networks (CNN) were used, taking advantage of the transfer learning technique (10). For ERG and eye fundus images, the following CNNs were used: ResNet-50, MobileNet-V2, and DenseNet-201. For the last two models, fine-tuning was also used to add four dense layers and two layers with LSTM networks at the end of the models. OpticNet-71 was used with OCT images and a fine-tuned version of DenseNet-201.

The bundles of non-evoked ERG-derived spectrogram and scalogram, fundus, and OCT images from the same patient were randomly assigned to training or test sets. A distribution percentage of 80 % training and 20 % testing was used.

To verify the possibility that the ResNet-50 and DenseNet-201 models’ performance is influenced by the presence of DR, we reclassified the wavelet scalogram images used for training and testing into images with (n = 293) or without some degree of DR (n = 896). To balance classes, scalograms from cases with DR were subjected to data augmentation, as described in the Methods, leading to the generation of 879 scalograms with some degree of DR. Data were randomly divided into training (80 %) and test (20 %), and the ResNet-50 and DenseNet-201 models were trained with the same parameters as described in the Methods, to compare the results between the binary classification of patients with and without DME, with the one of patients with and without DR. Validation was done using the previously used 164 images, though they were recategorized into those with some degree of DR (n = 77) and those without DR (n = 87). **Table 2** summarizes the metric performance of the models trained to distinguish DR and no DR classes. All metrics exceeded 0.60, unlike the values obtained for the validation of the DME class prediction, indicating that the concomitant presence of DR with DME takes part into the poor performance of our models.

**Table 2.** Comparative table between the validations done for EMD classification and DR classification. In general, the metrics obtained in validation with the same models, trained during the same number of epochs, are better in validation than those of DR classification. All the metrics are above 0.60 and Cohen’s Kappa is also improved.

| METRICS | MODELS |  |  |  |
| --- | --- | --- | --- | --- |
|  | DME vs No DME |  | DR vs No DR |  |
|  | ResNet-50 | DenseNet-201 | ResNet-50 | DenseNet-201 |
| **Precision** | 0.38 | 0.34 | 0.67 | 0.68 |
| **Accuracy** | 0.68 | 0.65 | 0.65 | 0.67 |
| **Sensitivity** | 0.36 | 0.40 | 0.66 | 0.70 |
| **Specificity** | 0.80 | 0.73 | 0.63 | 0.64 |
| **F1-Score** | 0.37 | 0.37 | 0.67 | 0.69 |
| **Cohen's Kappa** | 0.15 | 0.13 | 0.30 | 0.34 |

### Hybrid model

The best-performing models for each data type (OpticNet-71 for OCT, DenseNet-201 fine-tuning model trained with scalogram images, and DenseNet201-fine-tuning model for fundus images) were chosen to create a hybrid model. Predictions were obtained for each model using the 164 validation images, and the resulting prediction matrices were then multiplied by a weight factor (**Figure 1**). Input data (OCT, eye fundus, and scalogram images) came from the same eye of an individual and were taken the same day. To find the best weight values, all possible value combinations were tested. The weight distribution was selected based on the combination that yielded the best results for the hybrid model (**Figure 2**).

### Algorithm performance

Models with a loss below 0.4 and accuracy above 0.8 during training and testing were selected (**Table 3**). To evaluate the performance of the models, 164 images from OCT, eye fundus, and ERG-derived scalograms and spectrograms alien to training and test phases were used: 42 images with DME and 122 without DME in total, taken from 97 different subjects. The following metrics were calculated for each model: precision, sensitivity, specificity, accuracy, F1-score, the area under the curve (AUC) of the Receiver Operating Characteristic (ROC) curve, and Cohen’s Kappa (28).

**Table 3.** Models’ characteristics during training and testing, including the number of epochs for training and both the accuracy and loss after testing. OCT, optical coherence tomography images; FFT, FFT-derived spectrograms from basal ERGs; Wavelet, Wavelet transform-derived spectrograms from basal ERGs.

|  |  |  | Training |  | Test |  |
| --- | --- | --- | --- | --- | --- | --- |
| Model | Input | # Epochs | Accuracy | Loss | Accuracy | Loss |
| OpticNet-71 | OCT | 20 | 0.98 | 0.05 | 0.99 | 0.01 |
| Finely-tuned DenseNet-201 | OCT | 100 | 0.97 | 0.06 | 0.94 | 0.18 |
| ResNet-50 | Eye fundus | 50 | 1.00 | 0.01 | 0.99 | 0.03 |
| Finely-tuned MobileNet-V2 | Eye fundus | 100 | 0.96 | 0.10 | 0.99 | 0.03 |
| Finely-tuned DenseNet-201 | Eye fundus | 100 | 0.96 | 0.10 | 0.96 | 0.05 |
| ResNet-50 (FFT) | FFT | 50 | 0.94 | 0.18 | 0.93 | 0.16 |
| Finely-tuned DenseNet-201 | FFT | 200 | 0.84 | 0.37 | 0.94 | 0.18 |
| ResNet-50 | Wavelet | 50 | 0.98 | 0.10 | 0.96 | 0.16 |
| Finely-tuned DenseNet-201 | Wavelet | 200 | 0.88 | 0.27 | 0.95 | 0.15 |

### Code availability

To facilitate the reproducibility of our data analyses, the Python code and documentation for the analysis are available online (https://github.com/Traslational-Visual-Health-Laboratory).

## Data Availability

The Python code and documentation for the analysis are available online (https://github.com/Traslational-Visual-Health-Laboratory).

## Acknowledgements

We thank all volunteers under the care of the APEC, IMO, ENES León and INDEREB, as well as the many physicians and technicians who worked on data collection.

## Supporting information

**S1 figure**. Performance of all OCT models. ROC curves and confusion matrixes for the **(A)** OpticNet-71 model validated with n = 1,229 cases (n = 212 with DME and n = 1,017 without DME), **(B)** the Finely-tuned DenseNet-201 model validated with n = 1229 (n = 212 with DME and n = 1,017 without DME), **(C)** the OpticNet-71 model validated with n = 164 (n = 42 with DME and n = 122 without DME), and **(D)** the finely-tuned DenseNet-201 model validated with n = 1,229 (n = 212 with DME and n = 1017 without DME). **(D)** Summary of all above-models’ performance metrics.

**S2 figure**. Performance of the Eye Fundus-based model. ROC curves, confusion matrixes, and performance metrics for the **(A)** ResNet-50, **(B)** finely-tuned MobileNet-V2, and **(C)** finely-tuned DenseNet-201 models. The validation dataset included a total of 164 eye fundus images (n = 42 with DME and n = 122 without DME).

## Notes

### Competing Interest Statement

The authors have declared no competing interest.

### Funding Statement

J.A.H.C. is a Master student from the Programa de Maestría en Ciencias (Neurobiología), Universidad Nacional Autónoma de México (UNAM) and received a fellowship from the National Council of Science and Technology of Mexico (CONACYT CVU 1146197). This study was supported by the UNAM-DGAPA grant IN205420 (ST), IN212823 (ST), CONACYT 299625 (ST), and CONACYT CF-2019-1759 (ST) grants. The funders had no role in study design, data collection and analysis, decision to publish, or preparation of the manuscript.

